# Cathepsins and Glaucoma: Genetic Evidence from a Mendelian Randomization Approach

**DOI:** 10.1101/2025.07.20.25331784

**Authors:** Zhixiang Hua, Xiaoxiao Chen, Jin Yang, Xiaobo Yu

**Affiliations:** Department of Ophthalmology and the Eye Institute, Eye and Ear, Nose, and Throat Hospital, Fudan University, Shanghai 200000, China; Key NHC Key Laboratory of Myopia (Fudan University), Laboratory of Myopia, Chinese Academy of Medical Sciences, Shanghai 200031, China; Shanghai Key Laboratory of Visual Impairment and Restoration, Shanghai 200000, China

**Keywords:** Cathepsins, Glaucoma, Mendelian Randomization

## Abstract

**Background:** Glaucoma is a progressive optic neuropathy primarily associated with elevated intraocular pressure (IOP) and characterized by optic nerve damage. Despite numerous risk factors, including high IOP, the molecular mechanisms underlying glaucoma remain unclear. The cathepsin family, a group of lysosomal proteases, plays a critical role in various physiological and pathological processes. This study investigates the causal relationship between cathepsins and glaucoma using Mendelian Randomization (MR).

**Methods:** This two-sample MR study evaluates the causal relationship between nine cathepsins and glaucoma subtypes using genetic data from the INTERVAL study and the FinnGen consortium. The primary MR analysis used the Inverse Variance Weighting (IVW) method, with supplementary analyses including MR-Egger and Weighted Median methods. Reverse MR and multivariate MR analyses were also performed.

**Results:** Elevated levels of cathepsin F significantly decreased the risk of primary angle-closure glaucoma (PACG) (OR = 0.815, p = 0.005). Reverse MR analyses indicated that primary open-angle glaucoma (POAG) might reduce cathepsin F levels (OR = 0.949, p = 0.010). Multivariate MR analysis showed significant associations between specific cathepsins and glaucoma subtypes, including cathepsin F reducing the risk of PACG and cathepsin S reducing the risk of total glaucoma.

**Conclusion:** This study provides evidence of a causal relationship between cathepsin levels and glaucoma subtypes, particularly highlighting the protective role of cathepsin F against PACG. These research findings offer insights into potential therapeutic targets for glaucoma, with the elucidation of their deeper mechanisms awaiting further investigation.

## 1. Introduction

Glaucoma is a group of progressive optic neuropathy characterized by elevated intraocular pressure and optic nerve head damage.^[1]^ In 2040, the number of people aged 40–80 years affected by glaucoma worldwide was estimated to be 112 million.^[2]^ Based on the structural characteristics of the anterior chamber angle, glaucoma is categorized into two types: primary open-angle glaucoma (POAG) and angle-closure glaucoma (PACG). It is generally assumed that intraocular pressure (IOP) is the primary modifiable risk factor while progression usually stops if IOP is lowered by 30–50% from baseline which leading to damage to the optic nerve characteristic of glaucoma.^[3]^ Besides raised IOP, low ocular perfusion pressure, older age, a positive family history, high myopia also account for both development and progression of glaucoma.^[4]^ The molecular mechanism of POAG and PACG remains complicated and not well understood.

The human genome is meticulously programmed to encode a spectrum of proteolytic enzymes, with an estimated 500 to 600 proteases contributing to a vast array of biological functions.^[5]^ The cathepsin family is characterized by the presence of lysosomal proteases, which include aspartyl and cysteine proteases, as well as neutrophilic proteases.^[6]^ To date, eleven human cysteine cathepsins (B, C, H, F, K, L, O, S, L2/V, W, and Z) have been identified in mammalian cells. These entities play a pivotal role in a variety of critical physiological and pathological mechanisms, encompassing processes like proteolysis, autophagy, antigen presentation, the activation of immune cells, and the transmission of cellular signals.^[7]^ As we known, cathepsin are found in various location within the ocular tissures, including three layers of the cornea, the retinal pigment epithelial, the choroid and optic nerve.^[8–11]^ However, the relationship between cathepsins and glaucomatous optic nerve damage still remains unknown.

Mendelian randomization (MR) analysis is a robust method that incorporates genetic information into traditional epidemiologic methods as an alternative to RCT. MR leverages genetic variations as natural experiments to deduce the causal links between biological elements and diseases. By assessing the genotype distribution, which is established prior to disease onset, MR mitigates the influence of confounders and the issue of reverse causation.^[12]^ It provides a powerful tool for uncovering novel insights and delivering robust evidence of causal relationships among variables. MR has been instrumental in establishing the causal impacts of psychiatric disorders, diet-derived antioxidants, sleep behaviors and glaucoma.^[13–15]^ Utilizing the MR methodology, this study aimed to ascertain the causal interplay between cathepsins and various glaucoma subtypes.

## 2. Materials and Methods

### 2.1 Study Design

This investigation assessed the causal link between nine cathepsins and glaucoma susceptibility through a two-sample Mendelian Randomization framework, designating cathepsins as the exposure and glaucoma as the outcome.

Subsequently, reverse and multivariate MR analyses were executed, employing identical parameters and datasets as the initial forward MR analysis. The MR framework relied on three core assumptions: (1) Relevance: Selected single nucleotide polymorphisms (SNPs) must exhibit a strong association with levels of cathepsins (exposure). (2) Independence: SNPs should not correlate with potential confounding factors influencing both cathepsin levels and glaucoma risk. (3) Exclusion Restriction: SNPs must exert their influence on glaucoma risk solely via their effect on cathepsin levels, thus precluding genetic pleiotropy.The flow chart of this study is shown in Fig. 1.

**Figure 1.**
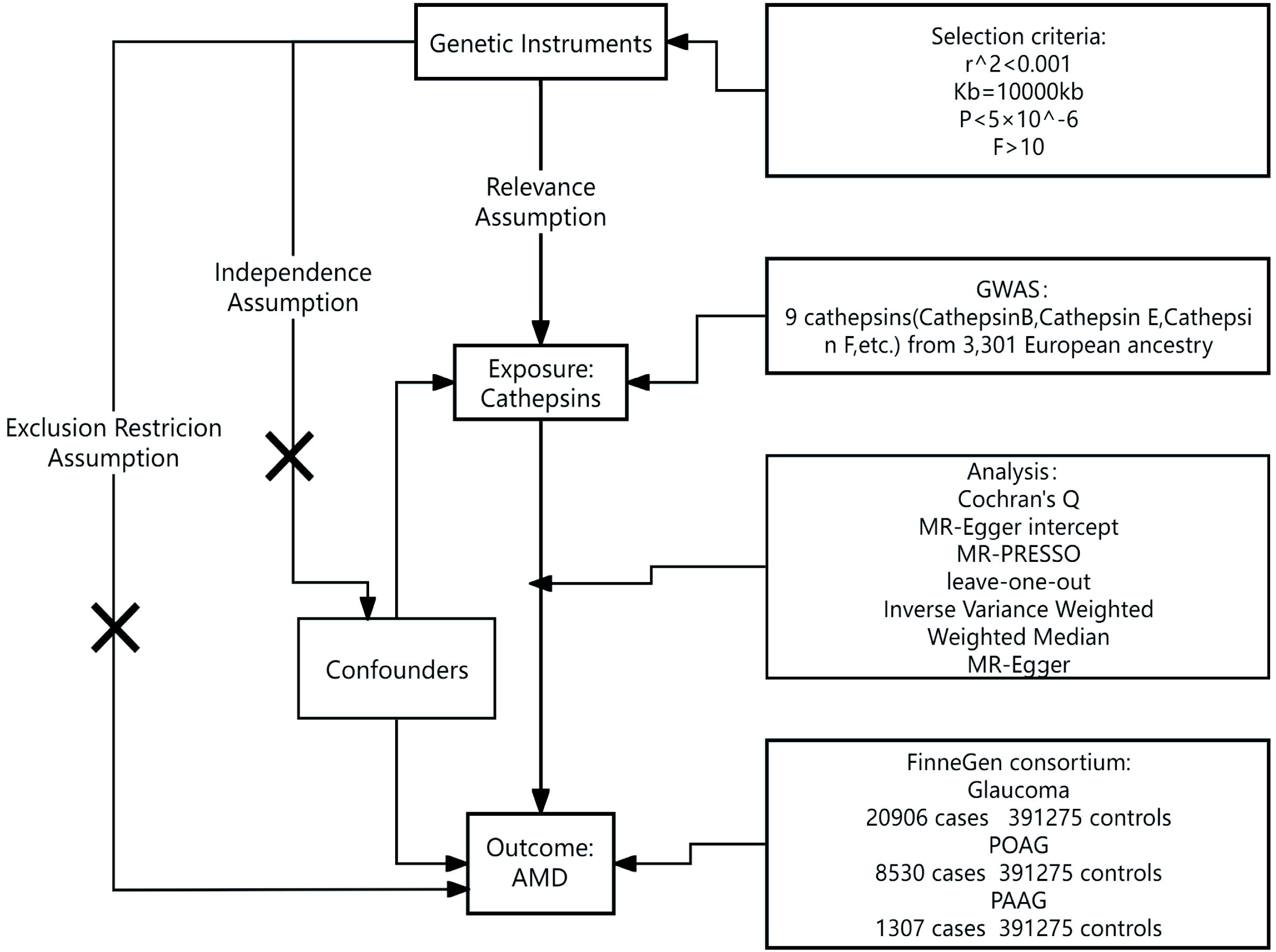
Flowchart of this MR analysis study design.

### 2.2 Data Sources

For the Mendelian randomization analysis, genetic instruments reflecting nine cathepsin levels were obtained from the INTERVAL study, which included individuals of European ancestry. Public access to the data is available through the GWAS database website (https://gwas.mrcieu.ac.uk.).^[16]^ The genetic data for glaucoma were sourced from the FinnGen consortium database (DF10).^[17]^ This extensive database comprises 20,906 cases of glaucoma, accompanied by 391,275 control individuals, ensuring a robust dataset for thorough analysis. Within this dataset, the POAG (strict) subset encompasses 8,530 cases and is complemented by 391,275 controls. Additionally, the PACG data consist of 1,307 cases and 391,275 controls. Together, these comprehensive datasets provide a valuable resource for investigating the genetic factors associated with different forms of glaucoma in a large population. Further specifics and data details are available via the official FinnGen website (https://www.finngen.fi/en/).

Participants in both contributing studies provided informed consent, and all research protocols received approval from their respective institutional ethics committees. The use of distinct GWAS databases for genetic samples aimed to mitigate overlap and bias, thereby obviating the need for further ethical clearances for this MR investigation.

### 2.3 Selection of Genetic Instruments

To precisely ascertain the causal link between cathepsins and glaucoma, a meticulous process was undertaken to select single nucleotide polymorphisms (SNPs) as instrumental variables. To affirm the validity and accuracy of this relationship, rigorous steps were undertaken to identify and select the most suitable SNPs associated with the exposure factors. Given the limited pool of SNPs available for MR analysis, a significance threshold was set at P < 5×10^−6^ to identify SNPs that demonstrated strong associations with the exposures being studied. Furthermore, to mitigate linkage disequilibrium effects, an r^2^ threshold of 0.001 and a clumping window of 10,000 kilobases were implemented. A critical step involved verifying that the chosen SNPs were devoid of associations with confounding variables that could skew the observed exposure-outcome relationships. Each SNP was thus rigorously confirmed to influence the outcome solely through its effect on the exposure factors, thereby ensuring an unadulterated causal pathway within the analysis.

### 2.4 Statistical Methods

The primary assessment of causality in this study relied on the Inverse Variance Weighting (IVW) method, with a P-value threshold of 0.05 indicating statistical significance. Causal effects were quantified using odds ratios (ORs) and their 95% confidence intervals (CIs); an OR below 1 suggested a protective influence, while an OR exceeding 1 implied a risk factor. To affirm the robustness of the results, MR-Egger and Weighted Median methods served as supplementary analyses. Cochran’s Q test was employed to assess SNP heterogeneity, and significant heterogeneity led to the use of a random effects model. Leave-one-out analysis was conducted to exclude SNPs with extreme influences. The MR-Egger intercept and MR-PRESSO global test were used to detect and adjust for horizontal pleiotropy. This MR study also incorporated a reverse analysis to examine potential reverse causation, using the same GWAS datasets with glaucoma as the exposure and cathepsin levels as the outcomes. Additionally, multivariable MR was applied to determine the direct causal impacts of individual cathepsins on distinct glaucoma subtypes, accounting for simultaneous multiple exposures.

### 2.5 Software and Tools

All statistical analyses were conducted using the R software, with specialized packages such as TwoSampleMR and MR-PRESSO employed for Mendelian Randomization analyses. R language packages, including ggplot2 and forestplot, were utilized to generate scatter plots and forest plots.

## 3. Results

### 3.1 MR Analysis on Glaucoma

This study employed Two-Sample Mendelian Randomization (MR) analyses to investigate the effects of nine cathepsins (B, E, F, G, H, L2, O, S, and Z) on the overall risk and various histological subtypes of glaucoma, including total glaucoma, primary open-angle glaucoma (POAG), and primary angle-closure glaucoma (PACG). The Inverse Variance Weighted (IVW) method served as the primary analytical technique to uncover specific causal relationships. Notably, the univariable MR analysis revealed that elevated levels of cathepsin F significantly decreased the risk of PACG, with an odds ratio (OR) of 0.815 and a p-value of 0.005 (Table 1).

### 3.2 MR Sensitivity Analysis

To counteract potential pleiotropic influences not captured by the IVW method, supplementary sensitivity analyses, specifically MR-Egger and Weighted Median analyses, were performed. Cochran’s Q test indicated no significant heterogeneity among the single nucleotide polymorphisms (SNPs) (p > 0.05), and the MR-Egger intercepts showed no evidence of horizontal pleiotropy, confirming the reliability and consistency of our findings. The stability of these causal estimates was further reinforced by leave-one-out analysis. Visual representations of these analyses, as presented in Fig. 2, underscore the coherence of results across various MR methodologies.

**Figure 2.**
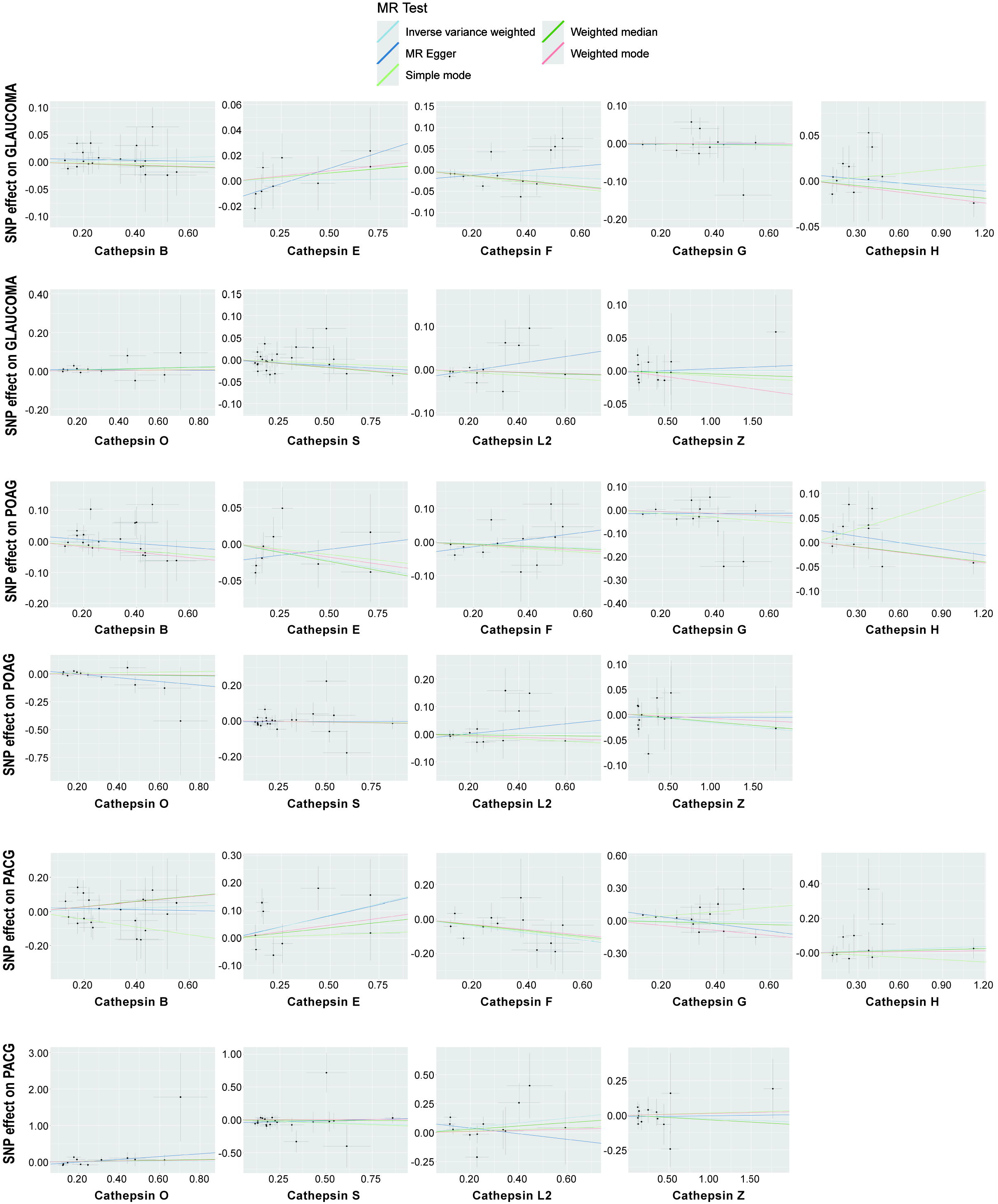
Scatter plots of MR analysis for cathepsins and glaucoma subtypes. Each point represents the effect of a single nucleotide polymorphism (SNP) on both the exposure and the outcome. Different colored regression lines indicate causal effects estimated using various algorithms.

### 3.3 Reverse MR Analysis

Reverse MR analyses were conducted to explore the potential for reverse causality between glaucoma and cathepsin levels. The analyses did not reveal significant reverse causation between glaucoma and the levels of various cathepsins. However, an intriguing finding was that POAG might reduce levels of cathepsin F, supported by the IVW method with a statistically significant OR of 0.949 (95% CI = 0.913–0.987, p = 0.010), with no indication of pleiotropy in the tests. The results of this reverse MR analysis are summarized in Table 2, which details the effects of glaucoma and its subtypes on cathepsins.

A multivariable Mendelian Randomization analysis was performed to assess the genetic predisposition involving multiple cathepsins concerning the risk of different glaucoma subtypes. After adjusting for the influence of other cathepsins, several significant causal associations were observed. Specifically, elevated levels of cathepsin F were robustly associated with a decreased risk of PACG, with an OR of 0.769 (95% CI = 0.642–0.921, p = 0.004). Additionally, elevated levels of cathepsin S were significantly associated with a decreased risk of total glaucoma (OR = 0.960, 95% CI = 0.929–0.992, p = 0.014), and cathepsin F also showed a significant protective effect against total glaucoma (OR = 0.947, 95% CI = 0.906–0.991, p = 0.018). Conversely, in PACG, cathepsin E was found to significantly increase the risk (OR = 1.212, 95% CI = 1.036–1.418, p = 0.017). The absence of horizontal pleiotropy in these analyses, confirmed by MR-Egger intercept analysis, supports the specificity of the observed effects. The results of the multivariable MR analysis are visually represented in the forest plots shown in Fig. 3, with detailed ORs presented in Table 3.

**Figure 3.**
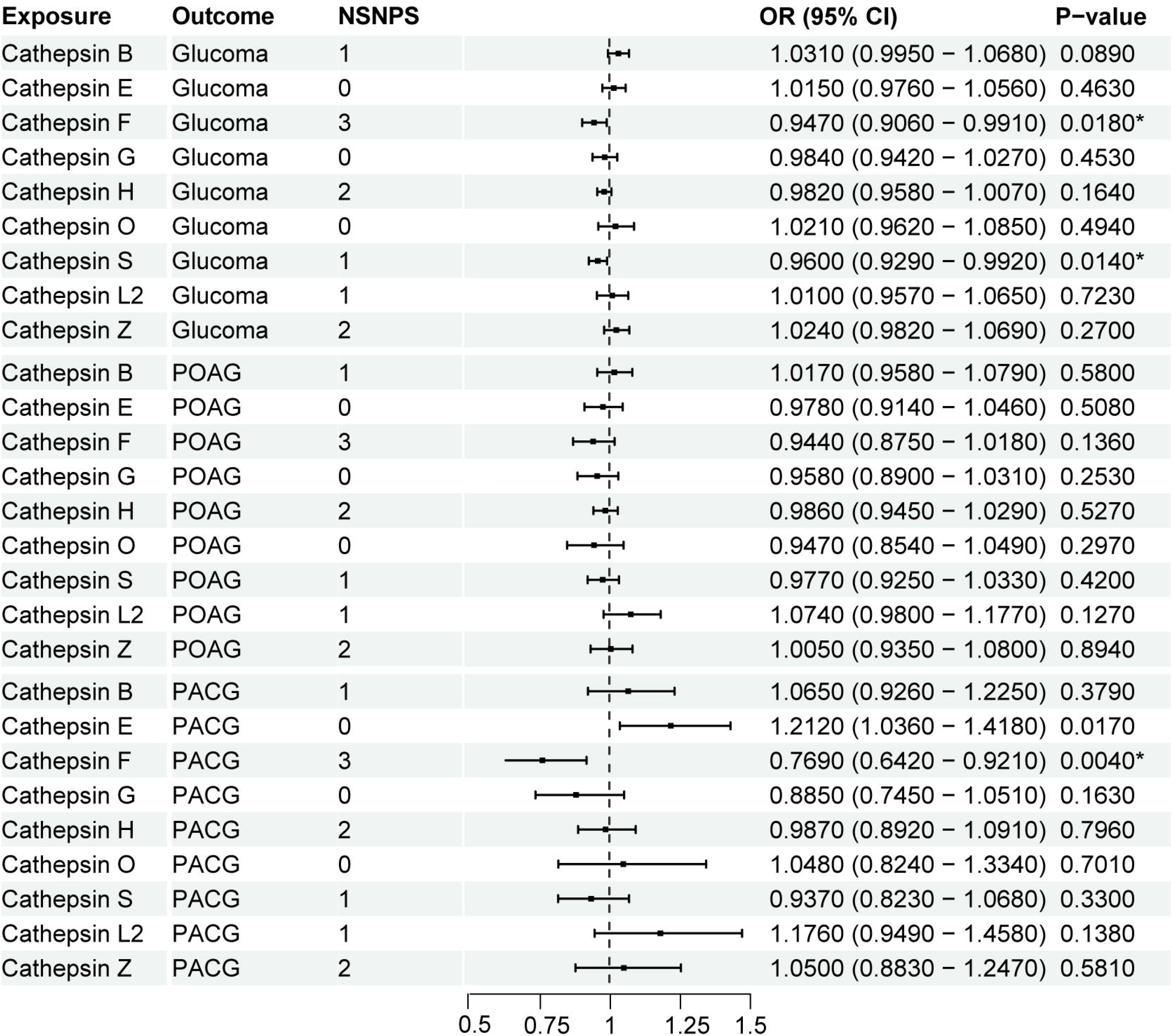
Linear forest plot illustrating the odds ratios (OR) from the multivariate Mendelian randomization (MR) analysis for the association between cathepsins and glaucoma. Each horizontal line represents an OR with its corresponding confidence interval, providing a clear visualization of effect sizes across different models or variables. * indicate statistically significant results with p-values less than 0.05.

## 4. Discussion

In this study, we used MR analyses to investigate the causal associations between cathepsins and glaucoma. Univariate MR analysis revealed that higher level of cathepsin F significantly reduced the risks of PACG. Reverse MR analysis reveals that POAG might reduce levels of cathepsin F suggesting the preventive role of cathepsin F in PACG and POAG from a genetic perspective.

PACG is distinguished by elevated IOP as a result of mechanical obstruction of the TM by either apposition of the peripheral iris to the TM or by a synechial closed angle. PACG can be classified according to the suddenness or timing of onset into acute PACG which defined as acute primary angle-closure (APAC) or chronic PACG.^[18]^ Anatomical features listing as mall cornea, thick lens, shallow anterior chamber, anterior lens position, and short axial length (AL) are anatomical features of PACG.^[19]^ The advent of advanced diagnostic modalities, including anterior segment optical coherence tomography (AS-OCT), enabling precise measurement of parameters involving iris thickness, iris volume and iris curvature.^[20]^

Genetically, the contribution of two single SNPs in PLEKHA7 and COL11A1 were confirmed in a large PACG cohort in China.^[21]^ Agreement with our result in this study, as molecules regulating ECM like COL11A1, cathepsin F was upregulated in PACG. Few literatures specifically report the role of cathepsin F in ophthalmic diseases. Saghizadeh etc. reported that silencing of cathepsin F at least partially normalized the patterns of several putative stem cell markers in diabetic organ-cultured human corneas.^[22]^ Other cathepsins such as cathepsin O was founded to be decreased in the aqueous humor of APAC patients.^[23]^ Additionally, the involvement of cathepsin D in Thiel-Behnke corneal dystrophy is attributed to its role in lysosomal dysfunction.^[24]^ Interestingly, cathepsin F inhibitor was found to be in the Coronavirus Disease 2019 (COVID-19) discovery phase.^[25]^ The incidence of APAC was proved to be increased while COVID-19 infection in different countries.^[26, 27]^ The mechanism of pathology and pathogenesis of COVID-19 related APAC have not been illustrated. The study on cathepsin F may become a breakthrough for COVID-related APAC.

Since POAG is defined as an acquired characteristic glaucomatous optic neuropathy associated with characteristic retinal ganglion cells loss and consequent irreversible visual field loss, the difference in the definition of PACG and POAG makes it difficult to compare prevalence and study risk factors in epidemiological glaucoma research.^[28]^ Although high IOP is the common risk factor for the progression in both PACG and POAG, Increased resistance to the outflow of aqueous humor (AH) is the main pathophysiological process in POAG. Resistance is derived of decreased outflow facility of AH either through the TM and Schlemm’s canal or via the uveoscleral outflow pathway.^[1]^ Functional failure of the TM tissue excessive ECM deposition impedes AH outflow leading to elevated IOP.^[29]^ Therefore, the breakthrough of research on lowering IOP in POAG has been concentrated on the TM with ECM. However, the exact mechanisms underlying TM cells dysfunction and ECM remodeling with IOP changes remain poorly understood

We firstly used the reverse MR analysis to find that POAG reduces cathepsin F level. Aging of the TM cells, or more precisely, that the production of reactive oxygen species (ROS) companied with the normal process of aging can be, at least partially, accounted for the loss in TM tissue in POAG.^[30]^ Cathepsin F is proved to be a potential marker for senescent human skin fibroblasts and keratinocytes associated with skin aging,^[31]^ of which maybe the key factor for the aging of TM cells in future studies. One of the potential cellular responses to ROS and oxidative damage is induction of autophagy.^[32]^ Autophagy is a process of intracellular self-digestion that facilitates the lysosomal degradation of long-lived proteins and organelles through the action of lysosomal hydrolases.^[33]^ Prolonged exposure of TM cells to oxidative stress, employed as an in vitro model of aging, has been observed to induce significant alterations within the lysosomal system. These alterations encompass an augmentation in lysosomal mass and the accumulation of autophagic vacuoles, as well as the deposition of oxidized materials within the lysosomes and the aggregation of damaged mitochondria. Concurrently, there is a notable reduction in the activity of cathepsin L, a key enzyme within the lysosomal system.^[34]^ The involvement of cathepsin F in other tissues were supported by Wang etc. in a glioblastoma xenograft mouse model therefore inhibiting autophagy levels in tumors.^[35]^

Several studies supported the other cathepsins involved in the progression of POAG via lysosomal dysfunction. Nettesheim etc. firstly reported a role of the cathepsin B in intracellular degradation of ECM components in TM cells. They found that degraded ECM products were associated with active cathepsin B in lysosomal compartments by regulating the expression of PAI-1 and TGFβ/Smad signaling.^[36]^ Besides, cathepsin B was also detected in AH of POAG patients. TM cells grown under oxidative stress conditions displayed, reduced lysosomal acidification and impaired cathepsin B proteolytic maturation, resulting in decreased autophagic flux.^[37]^

Although this MR analysis offers several strengths to complement traditional epidemiological research, our study has several limitations. Firstly, the data are derived from the INTERVAL study and the FinnGen consortium, which primarily involve participants of European descent. Thus, the results may not be generalizable to other populations. Additionally, although our findings have identified causal relationships between certain cathepsins and glaucoma, they have not provided detailed mechanistic insights into how cathepsins influence the progression of glaucoma. Further experimental studies are necessary to explore these mechanisms.

## Data Availability

All data produced are available online at:
https://gwas.mrcieu.ac.uk.
https://www.finngen.fi/en/

https://www.finngen.fi/en/

## Availability of Data and Materials

The data used in this study are sourced from publicly available databases. The relevant links to these databases have been provided in the main text of the manuscript. Additional data supporting the conclusions of this article are available from the corresponding author upon reasonable request..

## Author Contributions

Xiaobo Yu and Jin Yang designed the research; Zhixiang Hua collected and analyzed the data. Zhixiang Hua and Xiaoxiao Chen wrote the manuscript. Xiaobo Yu critically revised the manuscript. Zhixiang Hua and Xiaoxiao Chen contributed equally to this work.

## Ethics Approval and Consent to Participate

All participants in the studies provided written informed consent, and the research protocols were approved by the relevant institutional ethics committees. Due to the comprehensive nature of this approval process, no further ethical clearances were required for this Mendelian randomization study.

## Acknowledgment

We would like to extend our gratitude to the participants of the INTERVAL study and the FinnGen consortium, whose valuable contributions made this research possible. Their data were instrumental in advancing our understanding of the genetic factors associated with glaucoma.

## Abbreviations

IOP: Intraocular Pressure
PACG: Primary Angle-Closure Glaucoma
POAG: Primary Open-Angle Glaucoma
MR: Mendelian Randomization
IVW: Inverse Variance Weighted
OR: Odds Ratio
CI: Confidence Interval
SNPs: Single Nucleotide Polymorphisms
AS-OCT: anterior segment optical coherence tomography
AH: aqueous humor
AL: axial length
ECM: Extracellular Matrix
ROS: Reactive Oxygen Species
TM: Trabecular Meshwork
RCT: Randomized Controlled Trial

